# Bioimpedance Spectroscopy in Acute Decompensated Heart Failure: Insights on Fluid Status and Outcomes

**DOI:** 10.1101/2024.12.31.24319833

**Authors:** Shang-Feng Yang, I-Fan Liu, Hung-Yu Chang, Hsin-I Teng, Tien-Ping Tsao, Shin-Hung Tsai

## Abstract

**Background:** Acute decompensated heart failure (ADHF) is a leading cause of hospitalization, driven by fluid overload and associated with poor outcomes. Bioimpedance spectroscopy (BIS) provides a non-invasive method to assess fluid status, but its utility in predicting clinical outcomes, especially in chronic kidney disease (CKD) and heart failure with preserved ejection fraction (HFpEF), remains unclear.

**Methods:** This prospective study enrolled 157 patients hospitalized for ADHF. BIS was performed at admission and discharge to measure overhydration (OH) and the OH-to-extracellular water (OH/ECW) ratio. Patients were stratified into subgroups by CKD and heart failure with reduced ejection fraction (HFrEF). Outcomes included major adverse cardiovascular events (MACE) and heart failure hospitalization (HHF). Cox proportional hazards models, adjusted for age, sex, NT-proBNP, and ejection fraction, were used to evaluate the prognostic value of BIS parameters.

**Results:** Among the 157 patients, 72 (45.9%) had CKD, and 111 (70.7%) had HFrEF. CKD patients exhibited significantly higher OH and OH/ECW ratios at admission but showed no association with MACE or HHF. Greater reductions in OH/ECW were observed in HFrEF patients compared to non-HFrEF (P = 0.036), indicating better response to diuresis. However, these changes did not translate into improved clinical outcomes. Across all subgroups, OH, OH/ECW, and changes in these parameters were not significant predictors of MACE or HHF during the 10.7 ± 7.1-month follow-up.

**Conclusions:** BIS effectively monitors fluid status in ADHF patients, particularly in HFrEF. However, its prognostic utility in predicting outcomes in CKD and HFpEF subgroups is limited, warranting further investigation.

## Background

Acute decompensated heart failure (ADHF) is a leading cause of hospitalization, particularly in older individuals with cardiovascular disease. It is characterized by rapid or gradual worsening of heart failure (HF) symptoms, requiring immediate intervention to address fluid overload, pulmonary congestion, or inadequate perfusion.^1^ Despite advancements in treatment, ADHF is associated with high morbidity, frequent rehospitalizations, and poor outcomes. In-hospital mortality ranges from 4% to 10%, while 1-year post-discharge mortality can reach 25–30%, with readmission rates exceeding 45%.^2,3^

N-terminal pro-B-type natriuretic peptide (NT-proBNP) is a common biomarker for HF, indicating ventricular filling pressures and cardiac function.^4^ NT-proBNP levels are highly predictive of adverse outcomes in patients with ADHF.^5^ However, in chronic kidney disease (CKD) patients, its reliability decreases due to impaired renal clearance and chronic volume overload, which raise levels regardless of HF severity.^6^ Similarly, the NT-proBNP is less clear in HF with preserved left ventricular ejection fraction (LVEF).^7,8^ These limitations highlight the need for alternative approaches for accurate risk stratification and clinical implications.

Fluid overload is the primary cause of ADHF admissions and a major prognostic factor for post-discharge outcomes. Elevated congestion scores, incorporating orthopnea, jugular venous distension, and pedal edema, are strongly associated with increased mortality at 30 days and 1 year.^9^ However, traditional clinical assessments are often limited in accurately evaluating fluid status, as physical signs of edema do not reliably reflect intravascular volume.^10^ Bioimpedance analysis (BIA) and bioimpedance spectroscopy (BIS) are non-invasive, quantitative tools that offer objective assessments of fluid overload and decongestion.^11,12^

BIS provides precise measurements of overhydration (OH) and the OH-to-extracellular water (OH/ECW) ratio, enabling monitoring of fluid changes during hospitalization for ADHF. These metrics correlate with reductions in NT-proBNP, weight loss, and improvements in clinical parameters, making BIS a valuable tool for guiding decongestion strategies.^13,14^ Additionally, BIA enhances diagnostic accuracy when combined with biomarkers, particularly in the BNP ‘grey-zone,’ and provides prognostic value for 30-day outcomes like rehospitalization and mortality.^15,16^

This study aims to evaluate the utility of BIS in assessing fluid status and guiding treatment in ADHF patients, particularly those with CKD or normal ejection fraction. By comparing BIS-derived metrics with traditional markers such as NT-proBNP, the study seeks to determine whether BIS can predict clinical outcomes, including major adverse cardiovascular events and rehospitalization rates.

## Materials and Methods

### Study Design and Population

This prospective, observational study was conducted at Cheng Hsin General Hospital, Taipei, Taiwan, from January 2022 to December 2023. The study was designed to evaluate the utility of BIS in assessing and managing fluid overload in patients hospitalized for ADHF, with a focus on stratifying patients by CKD and heart failure with reduced ejection fraction (HFrEF). The study protocol was approved by the Institutional Review Board of Cheng Hsin General Hospital (IRB number: (898)110A-44), and informed consent was obtained from all participants.

### Inclusion and Exclusion Criteria

Patients aged 20 years or older, admitted with a primary diagnosis of ADHF, were eligible for inclusion. ADHF was defined as a worsening of heart failure symptoms such as dyspnea, orthopnea, and peripheral edema, accompanied by clinical evidence of fluid overload. Patients with CKD were defined by an estimated glomerular filtration rate (eGFR) <60 mL/min/1.73 m². Exclusion criteria included pregnancy, amputation, regular dialysis, acute myocardial infarction, severe valvular heart disease requiring surgical intervention, or those who were unable to undergo BIS measurements.

### Bioimpedance Spectroscopy

BIS was performed using the Fresenius Body Composition Monitor device, a non-invasive method that measures body fluid composition based on tissue resistance and reactance. BIS was conducted within 24 hours of admission and repeated at discharge. The key parameters measured were overhydration (OH, liter) and the ratio of overhydration to extracellular water (OH/ECW, %). Previous studies in CKD and ESRD populations have utilized these measurements to monitor volume status.^17^ These BIS measurements were used to track fluid status changes during hospitalization and were provided to the attending physicians for clinical management decisions.

### Data Collection

Demographic and clinical characteristics, including age, sex, body mass index (BMI), comorbidities (e.g., hypertension, diabetes), and LVEF, were collected at baseline. Laboratory values, including NT-proBNP, serum creatinine, and eGFR, were measured on admission. BIS measurements and the change in OH/ECW ratio between admission and discharge were also recorded. Patients were stratified by CKD and HFrEF status for subgroup analysis.

### Study Outcomes

The primary outcome of the study was the occurrence of major adverse cardiovascular events (MACE), defined as all-cause mortality, non-fatal myocardial infarction, non-fatal stroke, heart failure rehospitalization, or coronary revascularization. The secondary outcome was hospitalization for heart failure (HHF), defined as readmission due to worsening heart failure within the follow-up period. Patients were followed until death, December 31, 2023, or the last visit in our hospital, whichever came first.

### Statistical Analysis

Continuous variables were expressed as mean ± standard deviation (SD) for normally distributed data or as median with interquartile range (IQR) for non-normally distributed data, while categorical variables were presented as counts and percentages. Group comparisons between CKD and non-CKD or HFrEF and non-HFrEF were performed using the Student’s t-test, Mann-Whitney U test, or chi-square test, as appropriate. Cox proportional hazards regression models, adjusted for age, sex, LVEF, NT-proBNP, and baseline eGFR, assessed associations between OH, OH/ECW ratio changes, and clinical outcomes, with hazard ratios (HR) and 95% confidence intervals (CI) reported. Kaplan-Meier analysis examined cumulative events between groups with larger versus smaller OH/ECW reductions, with comparisons via log-rank tests. Statistical significance was set at P < 0.05, and analyses were conducted using SPSS version 25.0 (IBM SPSS Inc, Chicago, Illinois).

## Results

### Patient Demographics and Clinical Characteristics

A total of 157 patients hospitalized for acute heart failure (AHF) were enrolled in this study. Of the total cohort, 72 patients (45.9%) had CKD, and 111 patients (70.7%) had HFrEF. The demographic and clinical characteristics of these subgroups are summarized in **Tables 1 and 2**.

**Table 1.**
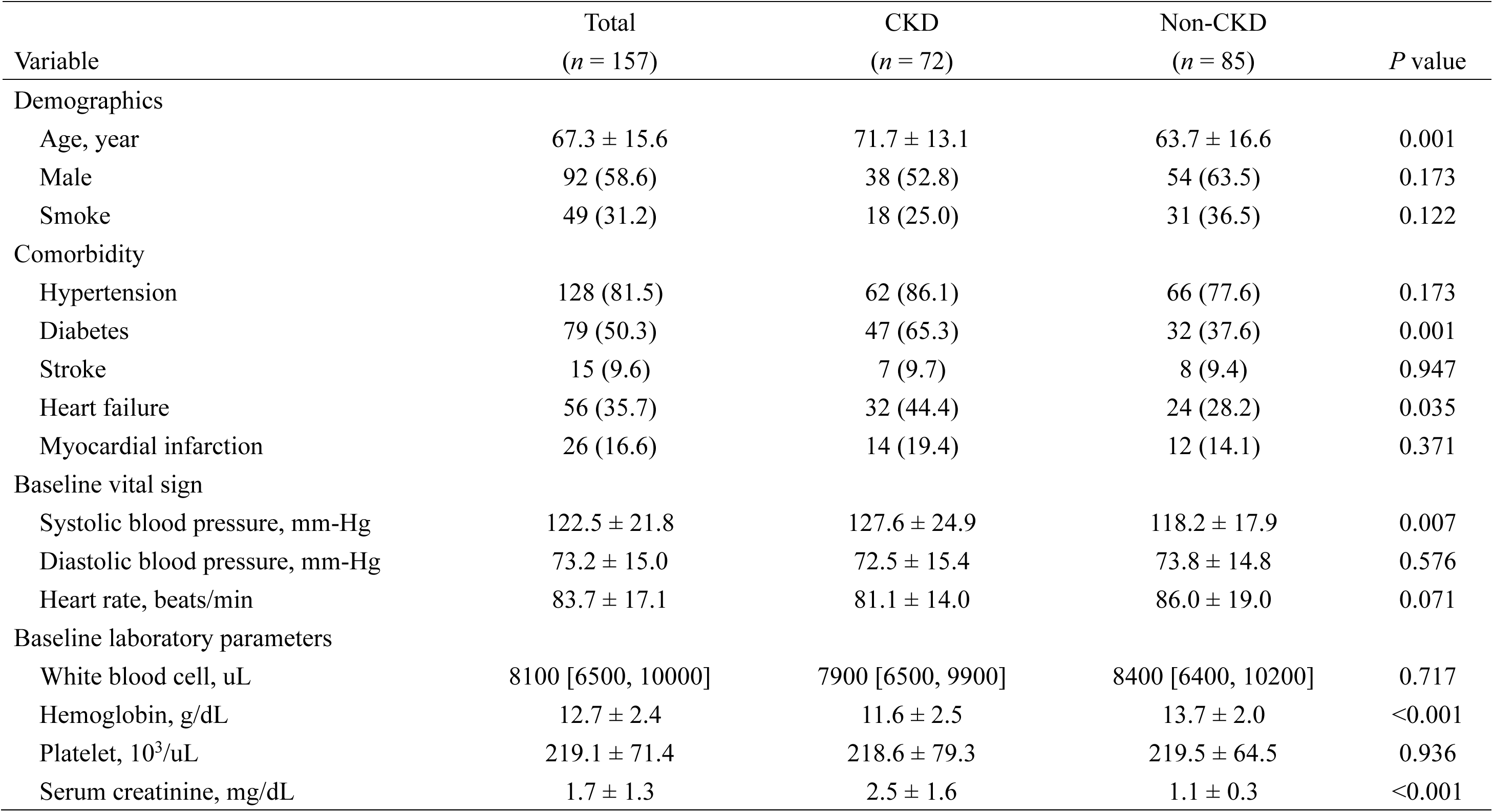
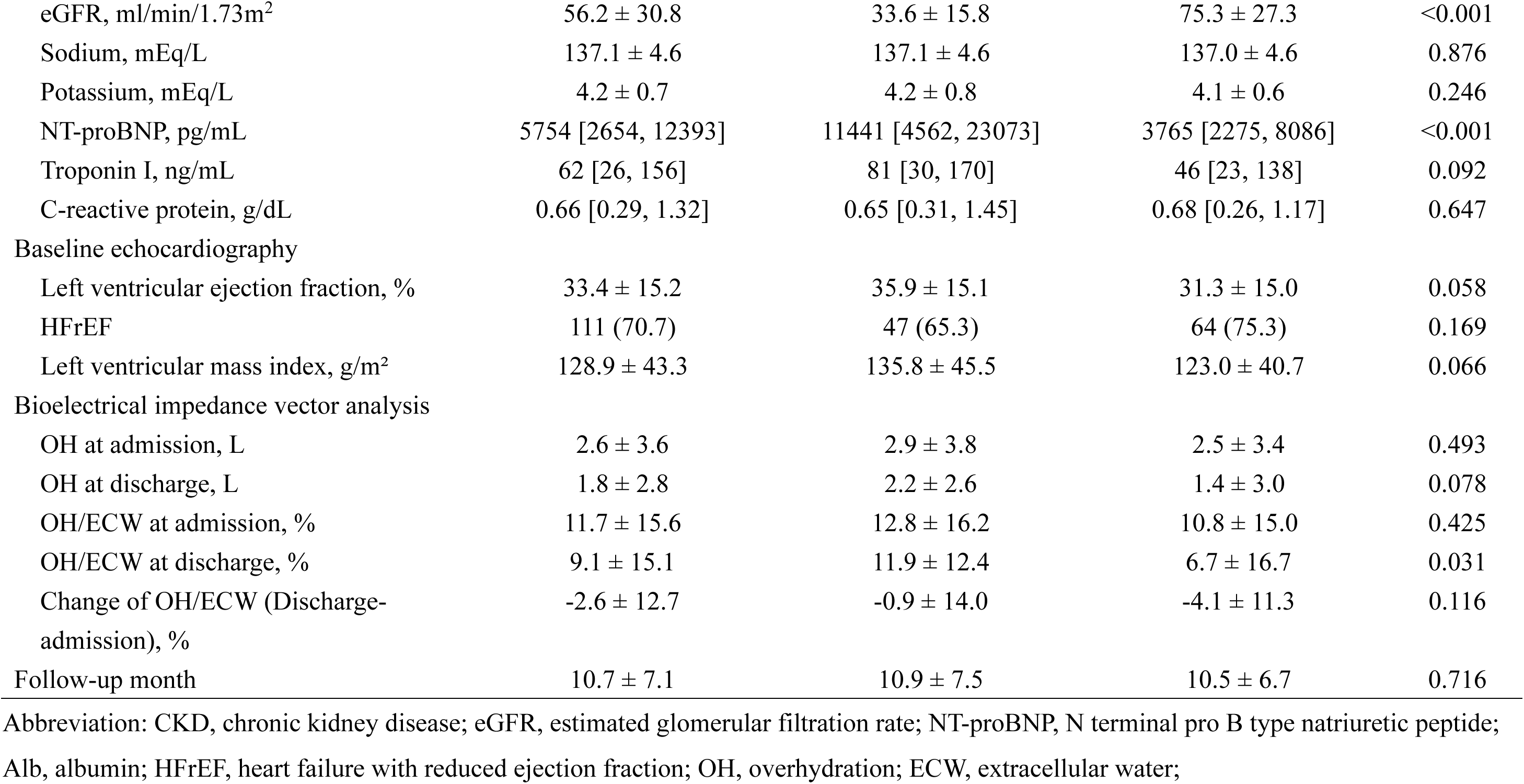
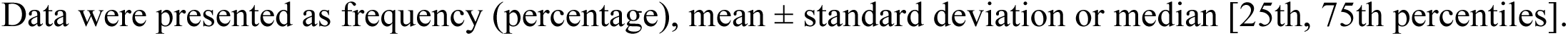
Demographic and clinical characteristics of patients with and without chronic kidney disease.

| Variable | Total<br>(*n* = 157) | CKD<br>(*n* = 72) | Non-CKD<br>(*n* = 85) | *P* value |
| --- | --- | --- | --- | --- |
| Demographics |  |  |  |  |
| Age, year | 67.3 ± 15.6 | 71.7 ± 13.1 | 63.7 ± 16.6 | 0.001 |
| Male | 92 (58.6) | 38 (52.8) | 54 (63.5) | 0.173 |
| Smoke | 49 (31.2) | 18 (25.0) | 31 (36.5) | 0.122 |
| Comorbidity |  |  |  |  |
| Hypertension | 128 (81.5) | 62 (86.1) | 66 (77.6) | 0.173 |
| Diabetes | 79 (50.3) | 47 (65.3) | 32 (37.6) | 0.001 |
| Stroke | 15 (9.6) | 7 (9.7) | 8 (9.4) | 0.947 |
| Heart failure | 56 (35.7) | 32 (44.4) | 24 (28.2) | 0.035 |
| Myocardial infarction | 26 (16.6) | 14 (19.4) | 12 (14.1) | 0.371 |
| Baseline vital sign |  |  |  |  |
| Systolic blood pressure, mm-Hg | 122.5 ± 21.8 | 127.6 ± 24.9 | 118.2 ± 17.9 | 0.007 |
| Diastolic blood pressure, mm-Hg | 73.2 ± 15.0 | 72.5 ± 15.4 | 73.8 ± 14.8 | 0.576 |
| Heart rate, beats/min | 83.7 ± 17.1 | 81.1 ± 14.0 | 86.0 ± 19.0 | 0.071 |
| Baseline laboratory parameters |  |  |  |  |
| White blood cell, uL | 8100 [6500, 10000] | 7900 [6500, 9900] | 8400 [6400, 10200] | 0.717 |
| Hemoglobin, g/dL | 12.7 ± 2.4 | 11.6 ± 2.5 | 13.7 ± 2.0 | <0.001 |
| Platelet, 10<sup>3</sup>/uL | 219.1 ± 71.4 | 218.6 ± 79.3 | 219.5 ± 64.5 | 0.936 |
| Serum creatinine, mg/dL | 1.7 ± 1.3 | 2.5 ± 1.6 | 1.1 ± 0.3 | <0.001 |
| eGFR, ml/min/1.73m<sup>2</sup> | 56.2 ± 30.8 | 33.6 ± 15.8 | 75.3 ± 27.3 | <0.001 |
| Sodium, mEq/L | 137.1 ± 4.6 | 137.1 ± 4.6 | 137.0 ± 4.6 | 0.876 |
| Potassium, mEq/L | 4.2 ± 0.7 | 4.2 ± 0.8 | 4.1 ± 0.6 | 0.246 |
| NT-proBNP, pg/mL | 5754 [2654, 12393] | 11441 [4562, 23073] | 3765 [2275, 8086] | <0.001 |
| Troponin I, ng/mL | 62 [26, 156] | 81 [30, 170] | 46 [23, 138] | 0.092 |
| C-reactive protein, g/dL | 0.66 [0.29, 1.32] | 0.65 [0.31, 1.45] | 0.68 [0.26, 1.17] | 0.647 |
| Baseline echocardiography |  |  |  |  |
| Left ventricular ejection fraction, % | 33.4 ± 15.2 | 35.9 ± 15.1 | 31.3 ± 15.0 | 0.058 |
| HFrEF | 111 (70.7) | 47 (65.3) | 64 (75.3) | 0.169 |
| Left ventricular mass index, g/m<sup>2</sup> | 128.9 ± 43.3 | 135.8 ± 45.5 | 123.0 ± 40.7 | 0.066 |
| Bioelectrical impedance vector analysis |  |  |  |  |
| OH at admission, L | 2.6 ± 3.6 | 2.9 ± 3.8 | 2.5 ± 3.4 | 0.493 |
| OH at discharge, L | 1.8 ± 2.8 | 2.2 ± 2.6 | 1.4 ± 3.0 | 0.078 |
| OH/ECW at admission, % | 11.7 ± 15.6 | 12.8 ± 16.2 | 10.8 ± 15.0 | 0.425 |
| OH/ECW at discharge, % | 9.1 ± 15.1 | 11.9 ± 12.4 | 6.7 ± 16.7 | 0.031 |
| Change of OH/ECW (Discharge-admission), % | -2.6 ± 12.7 | -0.9 ± 14.0 | -4.1 ± 11.3 | 0.116 |
| Follow-up month | 10.7 ± 7.1 | 10.9 ± 7.5 | 10.5 ± 6.7 | 0.716 |

Abbreviation: CKD, chronic kidney disease; eGFR, estimated glomerular filtration rate; NT-proBNP, N terminal pro B type natriuretic peptide; Alb, albumin; HFrEF, heart failure with reduced ejection fraction; OH, overhydration; ECW, extracellular water;
Data were presented as frequency (percentage), mean ± standard deviation or median [25th, 75th percentiles].

**Table 2.**
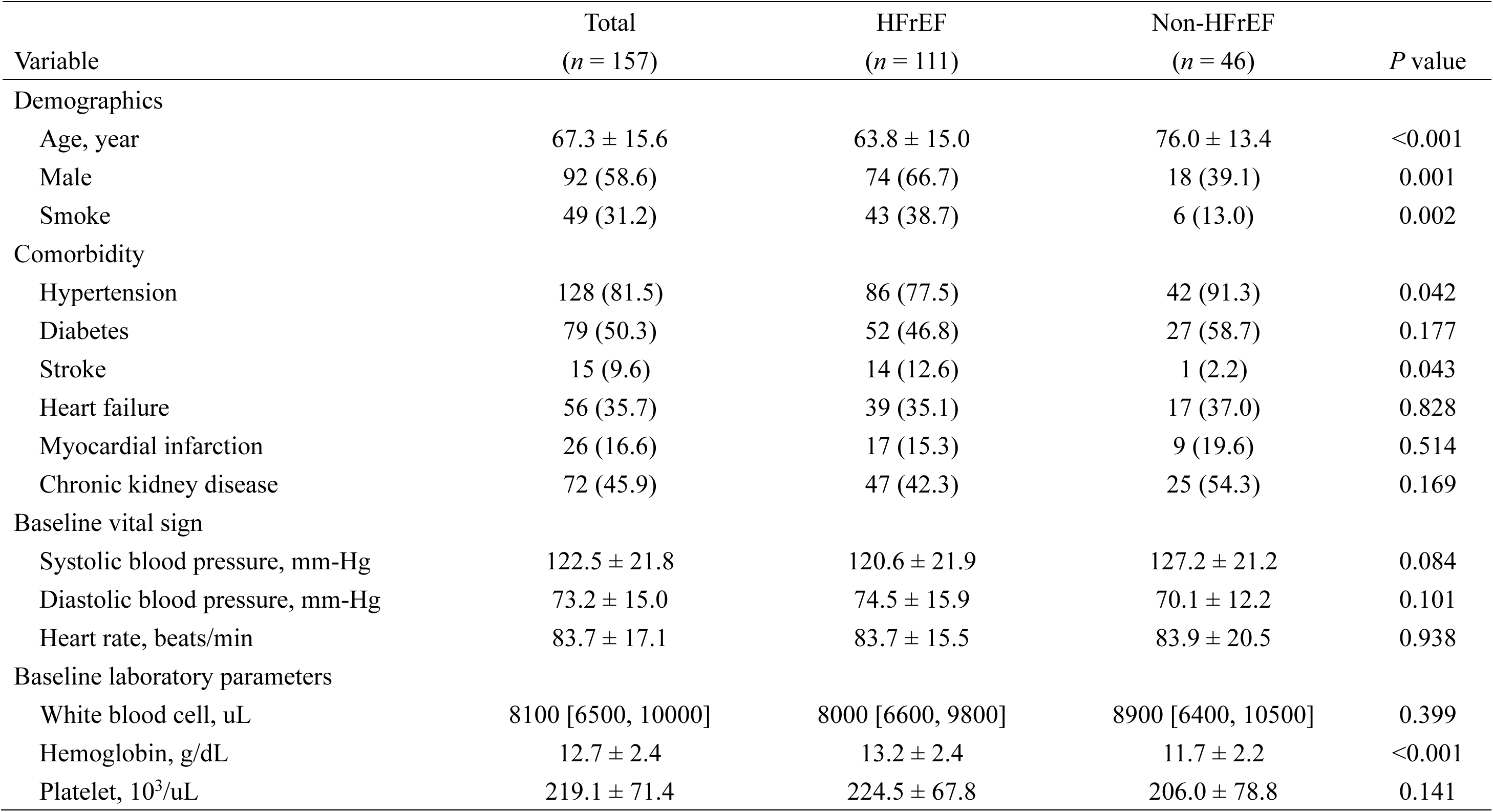
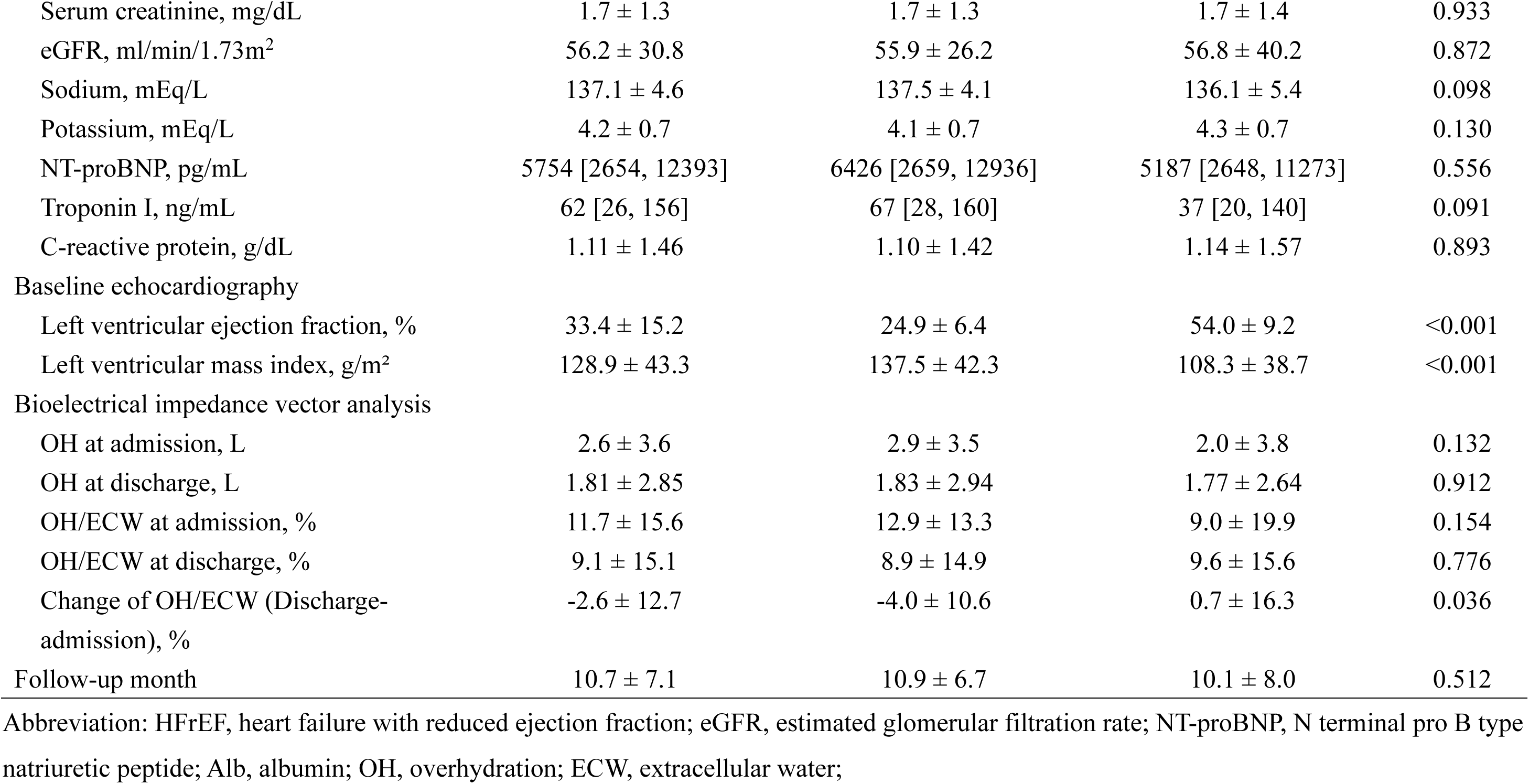
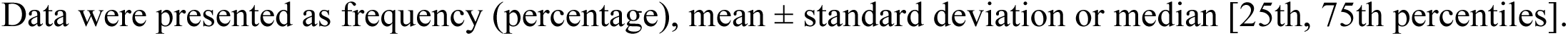
Demographic and clinical characteristics of patients with and without heart failure with reduced EF.

| Variable | Total<br>(*n* = 157) | HFrEF<br>(*n* = 111) | Non-HFrEF<br>(*n* = 46) | *P* value |
| --- | --- | --- | --- | --- |
| Demographics |  |  |  |  |
| Age, year | 67.3 ± 15.6 | 63.8 ± 15.0 | 76.0 ± 13.4 | <0.001 |
| Male | 92 (58.6) | 74 (66.7) | 18 (39.1) | 0.001 |
| Smoke | 49 (31.2) | 43 (38.7) | 6 (13.0) | 0.002 |
| Comorbidity |  |  |  |  |
| Hypertension | 128 (81.5) | 86 (77.5) | 42 (91.3) | 0.042 |
| Diabetes | 79 (50.3) | 52 (46.8) | 27 (58.7) | 0.177 |
| Stroke | 15 (9.6) | 14 (12.6) | 1 (2.2) | 0.043 |
| Heart failure | 56 (35.7) | 39 (35.1) | 17 (37.0) | 0.828 |
| Myocardial infarction | 26 (16.6) | 17 (15.3) | 9 (19.6) | 0.514 |
| Chronic kidney disease | 72 (45.9) | 47 (42.3) | 25 (54.3) | 0.169 |
| Baseline vital sign |  |  |  |  |
| Systolic blood pressure, mm-Hg | 122.5 ± 21.8 | 120.6 ± 21.9 | 127.2 ± 21.2 | 0.084 |
| Diastolic blood pressure, mm-Hg | 73.2 ± 15.0 | 74.5 ± 15.9 | 70.1 ± 12.2 | 0.101 |
| Heart rate, beats/min | 83.7 ± 17.1 | 83.7 ± 15.5 | 83.9 ± 20.5 | 0.938 |
| Baseline laboratory parameters |  |  |  |  |
| White blood cell, uL | 8100 [6500, 10000] | 8000 [6600, 9800] | 8900 [6400, 10500] | 0.399 |
| Hemoglobin, g/dL | 12.7 ± 2.4 | 13.2 ± 2.4 | 11.7 ± 2.2 | <0.001 |
| Platelet, 10<sup>3</sup>/uL | 219.1 ± 71.4 | 224.5 ± 67.8 | 206.0 ± 78.8 | 0.141 |
| Serum creatinine, mg/dL | 1.7 ± 1.3 | 1.7 ± 1.3 | 1.7 ± 1.4 | 0.933 |
| eGFR, ml/min/1.73m<sup>2</sup> | 56.2 ± 30.8 | 55.9 ± 26.2 | 56.8 ± 40.2 | 0.872 |
| Sodium, mEq/L | 137.1 ± 4.6 | 137.5 ± 4.1 | 136.1 ± 5.4 | 0.098 |
| Potassium, mEq/L | 4.2 ± 0.7 | 4.1 ± 0.7 | 4.3 ± 0.7 | 0.130 |
| NT-proBNP, pg/mL | 5754 [2654, 12393] | 6426 [2659, 12936] | 5187 [2648, 11273] | 0.556 |
| Troponin I, ng/mL | 62 [26, 156] | 67 [28, 160] | 37 [20, 140] | 0.091 |
| C-reactive protein, g/dL | 1.11 ± 1.46 | 1.10 ± 1.42 | 1.14 ± 1.57 | 0.893 |
| Baseline echocardiography |  |  |  |  |
| Left ventricular ejection fraction, % | 33.4 ± 15.2 | 24.9 ± 6.4 | 54.0 ± 9.2 | <0.001 |
| Left ventricular mass index, g/m<sup>2</sup> | 128.9 ± 43.3 | 137.5 ± 42.3 | 108.3 ± 38.7 | <0.001 |
| Bioelectrical impedance vector analysis |  |  |  |  |
| OH at admission, L | 2.6 ± 3.6 | 2.9 ± 3.5 | 2.0 ± 3.8 | 0.132 |
| OH at discharge, L | 1.81 ± 2.85 | 1.83 ± 2.94 | 1.77 ± 2.64 | 0.912 |
| OH/ECW at admission, % | 11.7 ± 15.6 | 12.9 ± 13.3 | 9.0 ± 19.9 | 0.154 |
| OH/ECW at discharge, % | 9.1 ± 15.1 | 8.9 ± 14.9 | 9.6 ± 15.6 | 0.776 |
| Change of OH/ECW (Discharge-admission), % | -2.6 ± 12.7 | -4.0 ± 10.6 | 0.7 ± 16.3 | 0.036 |
| Follow-up month | 10.7 ± 7.1 | 10.9 ± 6.7 | 10.1 ± 8.0 | 0.512 |

Abbreviation: HFrEF, heart failure with reduced ejection fraction; eGFR, estimated glomerular filtration rate; NT-proBNP, N terminal pro B type natriuretic peptide; Alb, albumin; OH, overhydration; ECW, extracellular water;
Data were presented as frequency (percentage), mean ± standard deviation or median [25th, 75th percentiles].

Patients with CKD were significantly older than those without CKD (71.7 ± 13.1 vs. 63.7 ± 16.6 years, *P* = 0.001). The CKD group also had a higher prevalence of diabetes (65.3% vs. 37.6%, *P* = 0.001) and heart failure (44.4% vs. 28.2%, *P* = 0.035). As expected, baseline renal function was worse in the CKD group, with a lower estimated glomerular filtration rate (eGFR) (33.6 ± 15.8 vs. 75.3 ± 27.3 mL/min/1.73m², *P* < 0.001) and higher serum creatinine levels (2.5 ± 1.6 vs. 1.1 ± 0.3 mg/dL, *P* < 0.001). Additionally, the CKD group had higher baseline NT-proBNP levels compared to the non-CKD group (11441 [4562–23073] vs. 3765 [2275–8086] pg/mL, *P* < 0.001).

In the HFrEF group, patients were younger than those in the non-HFrEF group (63.8 ± 15.0 vs. 76.0 ± 13.4 years, P < 0.001) and had a higher percentage of male patients (66.7% vs. 39.1%, P = 0.001). Smoking was also more prevalent in the HFrEF group (38.7% vs. 13.0%, P = 0.002). The prevalence of hypertension was significantly lower in the HFrEF group (77.5%) compared to the non-HFrEF group (91.3%, P = 0.042). Stroke was more common in the HFrEF group (9.6%) than in the non-HFrEF group (2.2%), with a P-value of 0.043. As expected, the LVEF was significantly lower in the HFrEF group compared to the non-HFrEF group (24.9 ± 6.4% vs. 54.0 ± 9.2%, P < 0.001), and the left ventricular mass index (LVMI) was higher in the HFrEF group (137.5 ± 42.3 vs. 108.3 ± 38.7 g/m², P < 0.001). Baseline hemoglobin levels were higher in the HFrEF group (13.2 ± 2.4 vs. 11.7 ± 2.2 g/dL, P < 0.001). Although baseline NT-proBNP levels were higher in the HFrEF group (6426 [2659–12936] vs. 5187 [2648–11273] pg/mL), this difference was not statistically significant (P = 0.556).

### Bioimpedance Spectroscopy Measurements in Subgroup of Interests

BIS was performed on admission and discharge to assess OH and its ratio to extracellular water (OH/ECW). The changes in these parameters were analyzed based on CKD and HFrEF status, as shown in **Table 1 and 2**.

In the CKD group, admission OH levels were slightly higher than in the non-CKD group (2.9 ± 3.8 vs. 2.5 ± 3.4 L, *P* = 0.493), but this difference was not statistically significant. At discharge, OH levels decreased in both groups, with a more pronounced reduction in the non-CKD group (2.2 ± 2.6 vs. 1.4 ± 3.0 L, *P* = 0.078). The OH/ECW ratio on admission was also higher in the CKD group but not significantly (12.8 ± 16.2 vs. 10.8 ± 15.0%, *P* = 0.425); however, at discharge, the ratio was significantly lower in the non-CKD group (11.9 ± 12.4 vs. 6.7 ± 16.7%, *P* = 0.031).

In the HFrEF group, OH levels at admission were higher than those in the non-HFrEF group (2.9 ± 3.5 vs. 2.0 ± 3.8 L, *P* = 0.132) but not significantly different, and discharge OH levels were comparable (1.83 ± 2.94 vs. 1.77 ± 2.64 L, *P* = 0.912). However, the change in the OH/ECW ratio from admission to discharge was significantly greater in the HFrEF group (−4.0 ± 10.6 vs. 0.7 ± 16.3%, *P* = 0.036), indicating a larger reduction in volume status.

### MACE and HHF in total participants

As shown in **Table 3**, 51 patients (32.5%) experienced MACE and 45 (28.7%) experienced HHF during an average follow-up period of 10.7 ± 7.1 months. For MACE, neither OH at admission nor at discharge demonstrated statistically significant predictive value across any model. In Model 3, adjusted for age, sex, prior heart failure hospitalization, LVEF, NT-proBNP levels, and eGFR, the hazard ratio (HR) for OH at admission was 0.993 (95% CI 0.976–1.011, P = 0.464), and the HR for OH at discharge was 0.988 (95% CI 0.968–1.009, P = 0.262). Similarly, changes in the OH/ECW ratio from admission to discharge were not significantly associated with the risk of MACE. Specifically, patients with smaller reductions in OH/ECW (≥-2.8%) had an HR of 1.15 (95% CI 0.65–2.04, P = 0.640) compared to those with larger reductions (<-2.8%).

**Table 3.**
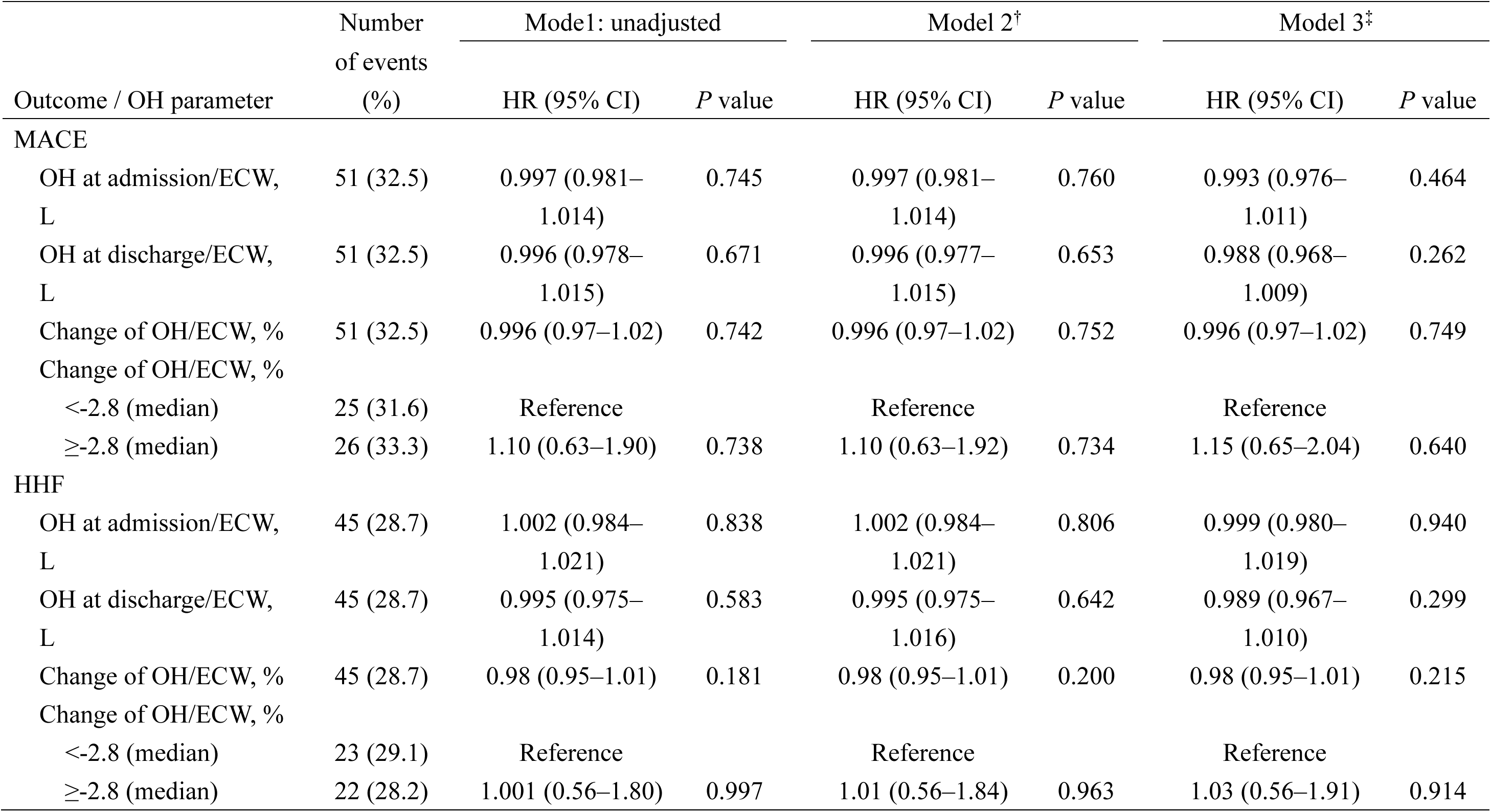
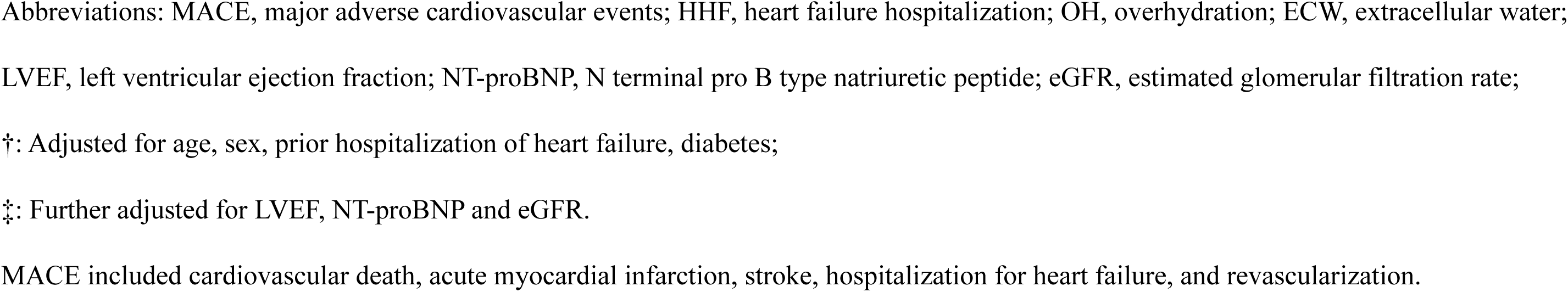
The association between overhydration (admission, discharge and change) and the risk of MACE and HHF.

|  | Number<br>of events | Model 1: unadjusted |  | Model 2<sup>†</sup> |  | Model 3<sup>‡</sup> |  |
| --- | --- | --- | --- | --- | --- | --- | --- |
| Outcome / OH parameter | (%) | HR (95% CI) | *P* value | HR (95% CI) | *P* value | HR (95% CI) | *P* value |
| MACE |  |  |  |  |  |  |  |
| OH at admission/ECW, L | 51 (32.5) | 0.997 (0.981–1.014) | 0.745 | 0.997 (0.981–1.014) | 0.760 | 0.993 (0.976–1.011) | 0.464 |
| OH at discharge/ECW, L | 51 (32.5) | 0.996 (0.978–1.015) | 0.671 | 0.996 (0.977–1.015) | 0.653 | 0.988 (0.968–1.009) | 0.262 |
| Change of OH/ECW, % | 51 (32.5) | 0.996 (0.97–1.02) | 0.742 | 0.996 (0.97–1.02) | 0.752 | 0.996 (0.97–1.02) | 0.749 |
| Change of OH/ECW, % |  |  |  |  |  |  |  |
| <-2.8 (median) | 25 (31.6) | Reference |  | Reference |  | Reference |  |
| ≥-2.8 (median) | 26 (33.3) | 1.10 (0.63–1.90) | 0.738 | 1.10 (0.63–1.92) | 0.734 | 1.15 (0.65–2.04) | 0.640 |
| HHF |  |  |  |  |  |  |  |
| OH at admission/ECW, L | 45 (28.7) | 1.002 (0.984–1.021) | 0.838 | 1.002 (0.984–1.021) | 0.806 | 0.999 (0.980–1.019) | 0.940 |
| OH at discharge/ECW, L | 45 (28.7) | 0.995 (0.975–1.014) | 0.583 | 0.995 (0.975–1.016) | 0.642 | 0.989 (0.967–1.010) | 0.299 |
| Change of OH/ECW, % | 45 (28.7) | 0.98 (0.95–1.01) | 0.181 | 0.98 (0.95–1.01) | 0.200 | 0.98 (0.95–1.01) | 0.215 |
| Change of OH/ECW, % |  |  |  |  |  |  |  |
| <-2.8 (median) | 23 (29.1) | Reference |  | Reference |  | Reference |  |
| ≥-2.8 (median) | 22 (28.2) | 1.001 (0.56–1.80) | 0.997 | 1.01 (0.56–1.84) | 0.963 | 1.03 (0.56–1.91) | 0.914 |

Abbreviations: MACE, major adverse cardiovascular events; HHF, heart failure hospitalization; OH, overhydration; ECW, extracellular water;
LVEF, left ventricular ejection fraction; NT-proBNP, N terminal pro B type natriuretic peptide; eGFR, estimated glomerular filtration rate;
†: Adjusted for age, sex, prior hospitalization of heart failure, diabetes;
‡: Further adjusted for LVEF, NT-proBNP and eGFR.
MACE included cardiovascular death, acute myocardial infarction, stroke, hospitalization for heart failure, and revascularization.

### Kaplan-Meier Survival Analysis in Subgroup of Interests

Kaplan-Meier survival curves were generated to assess the cumulative incidence of MACE and HHF in the study population, stratified by CKD (**Figures 1**) and HFrEF (**Figures 2**) status. The outcomes were compared between patients with greater and smaller reductions in OH/ECW within each subgroup, providing insights into the impact of fluid status changes on clinical events.

**Figure 1.**
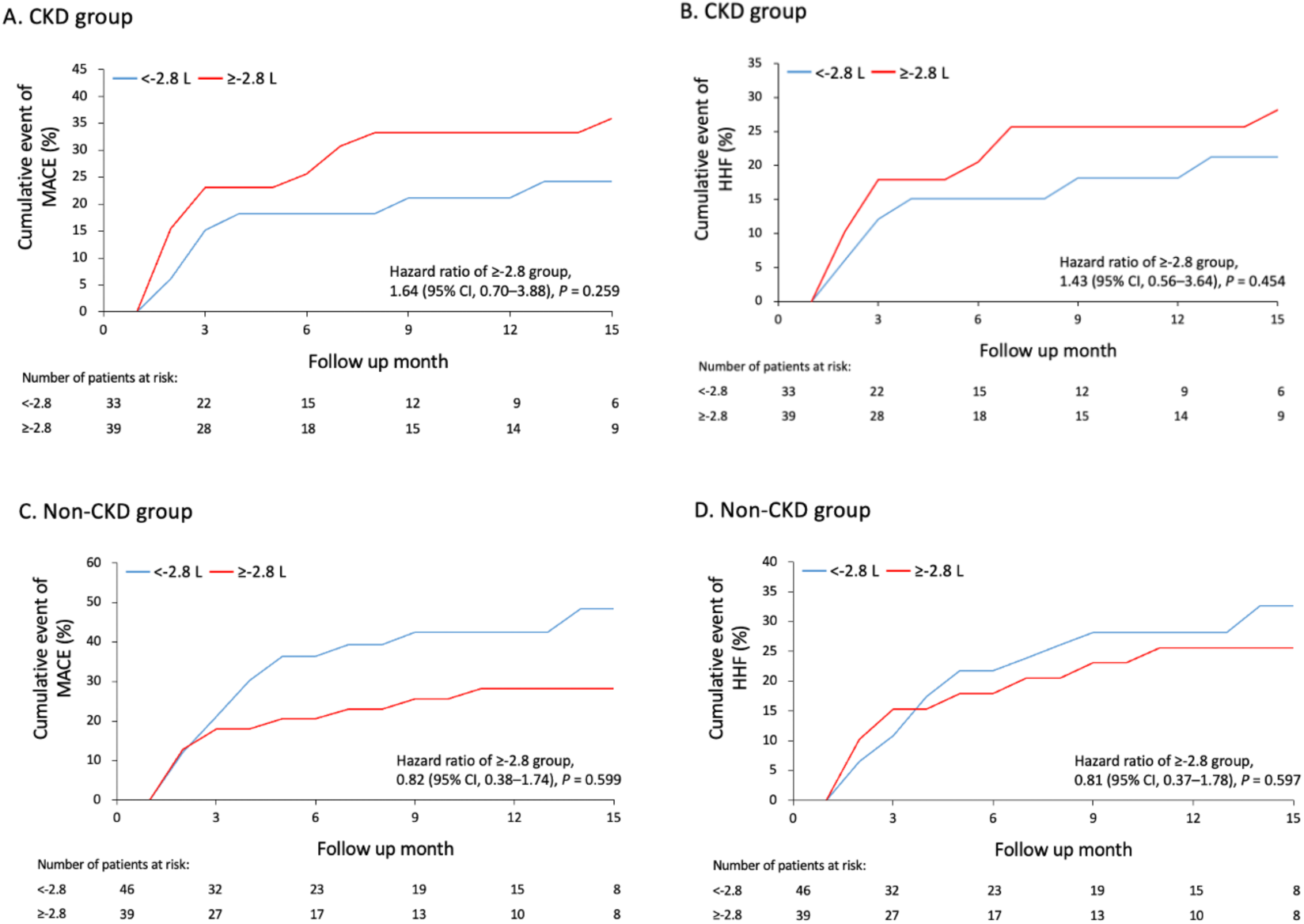
Kaplan-Meier Curves for MACE and HHF Based on OH/ECW Changes by CKD Status. The figure illustrates the cumulative incidence of MACE and HHF stratified by changes in the OH/ECW ratio (<-2.8 L vs. ≥-2.8 L) in patients with and without CKD. Abbreviations: OH/ECW, overhydration-to-extracellular water ratio; CKD, chronic kidney disease; MACE, major adverse cardiovascular events; HHF, heart failure hospitalization.

**Figure 2.**
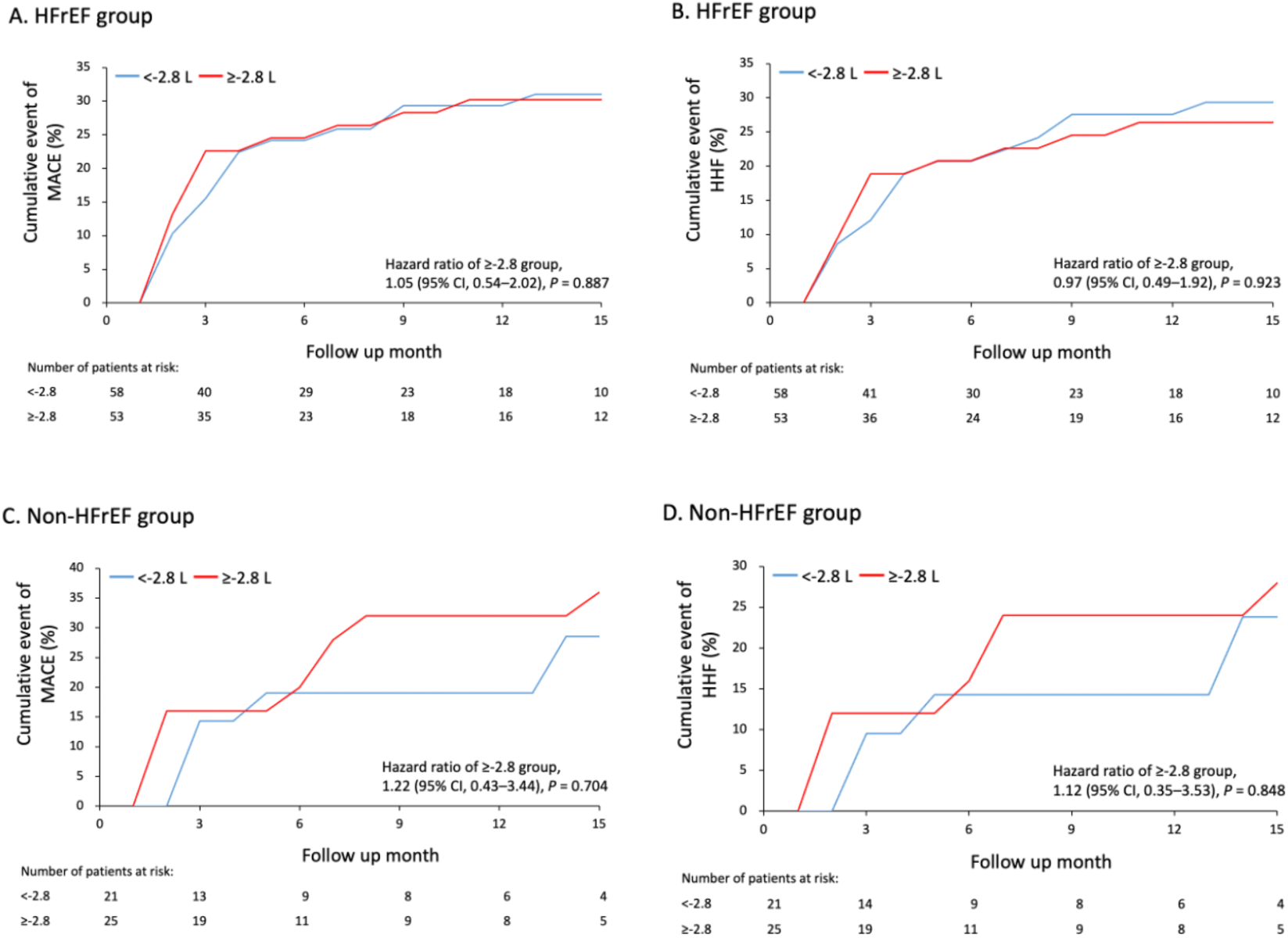
Kaplan-Meier Curves for MACE and HHF Based on OH/ECW Changes by HFrEF Status. This figure displays the cumulative incidence of MACE and HHF stratified by OH/ECW changes (<-2.8 L vs. ≥-2.8 L) in patients with HFrEF and non-HFrEF. Abbreviations: OH/ECW, overhydration-to-extracellular water ratio; HFrEF, heart failure with reduced ejection fraction; MACE, major adverse cardiovascular events; HHF, heart failure hospitalization.

In the CKD group (**Figure 1A and 1B**), patients with smaller reductions in OH/ECW (≥-2.8 L) showed no significant difference in the risk of MACE or HHF compared to those with larger reductions in OH/ECW, with HR of 1.64 (95% CI 0.70–3.88, P = 0.259) for MACE and 1.43 (95% CI 0.56–3.64, P = 0.454) for HHF. Similarly, in the non-CKD group (**Figure 1C and 1D**), no significant differences were observed, with HR of 0.82 (95% CI 0.38–1.74, P = 0.599) for MACE and 0.81 (95% CI 0.37–1.78, P = 0.597) for HHF. The cumulative incidence of MACE and HHF was also not significantly different in either the CKD or non-CKD groups based on OH/ECW reductions.

In patients with HFrEF (**Figure 2A-2B**), changes in the OH/ECW ratio did not significantly impact clinical outcomes. The HR for MACE was 1.05 (95% CI 0.54– 2.02, P = 0.887) and for HHF was 0.97 (95% CI 0.49–1.92, P = 0.923), showing no significant association between smaller OH/ECW reductions and higher risks. Similarly, in the non-HFrEF group (**Figure 2C-2D**), the HR for MACE was 1.22 (95% CI 0.43–3.44, P = 0.704) and for HHF was 1.12 (95% CI 0.35–3.53, P = 0.848), with no significant difference.

## Discussion

This study evaluated the utility of BIS in assessing fluid overload and predicting clinical outcomes in patients hospitalized for ADHF. Patients were stratified by CKD and HFrEF status to examine the role of BIS in different subpopulations. The findings indicated that OH values and changes in ratio of OH/ECW, as measured by BIS, did not significantly predict MACE or HHF in either CKD or non-HFrEF groups. However, BIS was effective in measuring fluid overload, and reductions in OH/ECW ratio were greater in patients with HFrEF compared to non-HFrEF patients.

BIS has been increasingly used in clinical settings to assess fluid status. BIS is a non-invasive tool that provides real-time data on body fluid compartments, which is particularly useful in managing conditions characterized by fluid overload, such as ADHF.^18,19^ In this study, BIS measurements were performed on admission and discharge to evaluate changes in fluid overload during hospitalization. Despite its ability to measure changes in fluid status, the overall impact of BIS measurements on clinical outcomes such as MACE and HHF was not significant in our population. This finding aligns with prior studies that have suggested that while BIS provides valuable insights into fluid overload, it may not directly predict adverse cardiovascular outcomes in all critically ill patients.^20^ Nonetheless, the information provided by BIS can still be valuable in guiding fluid management strategies during hospitalization, particularly in patients with complicated fluid balance issues.

Patients with CKD are particularly prone to volume overload, and BIS may be a valuable tool to guide management. Measures like phase angle and OH index predict adverse outcomes, including mortality and cardiovascular events, highlighting it’s prognostic utility in this population.^21,22^ Unlike NT-proBNP, which is often unreliable in CKD due to impaired renal clearance, BIS provides more accurate insights into fluid status, aiding in therapy optimization.^23^ In our study, BIS-derived OH and OH/ECW were higher in CKD patients compared to non-CKD patients, yet these metrics did not predict clinical outcomes. This underscores the need for larger studies to confirm the prognostic value of BIS in CKD patients.

Managing ADHF patients with different LVEF levels is complicated, as HFpEF patients generally exhibit lower NT-proBNP levels, indicating different underlying mechanisms than HFrEF.^24,25^ BIS may offer a more reliable alternative for evaluating volume status in this subgroup. Despite this potential, our study did not find significant differences in clinical outcomes related to changes in OH/ECW in non-HFrEF group, consistent with prior findings that fluid overload is less strongly correlated with adverse outcomes in HFpEF compared to HFrEF. These findings suggest that while BIS is useful for monitoring fluid status, its prognostic utility in non-HFrEF remains uncertain. Future studies should explore the integration of BIS with other diagnostic tools to optimize management in this population.

Notably, our study found a greater reduction in OH/ECW ratio in HFrEF patients compared to non-HFrEF patients, despite similar OH and OH/ECW ratios at admission and discharge. This suggests that ADHF in HFrEF may be more volume-dependent, while non-HFrEF could involve mechanisms such as fluid redistribution, anemia, arrhythmia, neurohormonal activation, or comorbidities like renal or pulmonary dysfunction.^26^ Besides, A prior study on volume status and central hemodynamics in HF patients showed elevated filling pressures even without hypervolemia, suggesting aggressive diuresis therapies may not always be suitable.^27^ Properly addressing underlying causes, especially in patients with preserved LVEF, is critical for effective treatment. Interestingly, a small pilot study reported greater total fluid loss and weight reduction in HFpEF patients compared to HFrEF during aggressive diuresis.^28^ The variations in volume changes among different etiologies of ADHF during active treatment warrant further detailed investigation.

This study has several limitations. First, it was conducted at a single center with a small sample size, limiting the generalizability of the findings. Second, while BIS provides a non-invasive method to assess fluid status, it may not fully capture the complexity of fluid dynamics in heart failure, especially in patients with comorbidities like CKD. The absence of a control group without BIS measurements prevents evaluation of whether BIS-guided management improves outcomes compared to traditional methods. Additionally, the follow-up period of 10.7 ± 7.1 months may not be sufficient to assess long-term outcomes, such as mortality. Lastly, reliance on OH/ECW as the primary marker of fluid overload may overlook other factors like tissue perfusion and organ congestion.

In conclusion, BIS is a reliable, non-invasive tool for assessing fluid overload in patients hospitalized for ADHF. While it did not predict outcomes such as MACE or HHF, it effectively measured changes in fluid status, particularly in HFrEF patients with larger OH/ECW reductions. The utility of BIS in CKD and HFpEF remains unclear, requiring further studies with larger cohorts and longer follow-ups to validate its prognostic value. Despite these limitations, BIS could still guide therapy in complex cases where traditional biomarkers are less reliable. Future research should explore integrating BIS with other hemodynamic and laboratory markers to optimize fluid management and outcomes in heart failure patients.

## Data Availability

The data that support the findings of this study are available from the corresponding author upon reasonable request. Due to patient confidentiality and ethical considerations, individual-level data cannot be shared publicly. Aggregate data, de-identified participant data, and additional methodological details can be provided to qualified researchers upon formal request and approval by the institutional review board.

## Acknowledgments

The authors would like to thank the study nurses (Chia-Chien Chang, Hsiu-Mien Chang and Yen-Lin Huang) who contributed to data collection.

